# Genomic analysis identifies risk factors in restless legs syndrome

**DOI:** 10.1101/2023.12.19.23300211

**Authors:** Fulya Akçimen, Ruth Chia, Sara Saez-Atienzar, Paola Ruffo, Memoona Rasheed, Jay P. Ross, Calwing Liao, Anindita Ray, Patrick A. Dion, Sonja W. Scholz, Guy A. Rouleau, Bryan J. Traynor

## Abstract

Restless legs syndrome (RLS) is a neurological condition that causes uncomfortable sensations in the legs and an irresistible urge to move them, typically during periods of rest. The genetic basis and pathophysiology of RLS are incompletely understood. Here, we present a whole-genome sequencing and genome-wide association meta-analysis of RLS cases (n = 9,851) and controls (n = 38,957) in three population-based biobanks (All of Us, Canadian Longitudinal Study on Aging, and CARTaGENE). Genome-wide association analysis identified nine independent risk loci, of which eight had been previously reported, and one was a novel risk locus (*LMX1B*, rs35196838, OR = 1.14, 95% CI = 1.09-1.19, *p*-value = 2.2 × 10^-9^). A genome-wide, gene-based common variant analysis identified *GLO1* as an additional risk gene (*p*-value = 8.45 × 10^-7^). Furthermore, a transcriptome-wide association study also identified *GLO1* and a previously unreported gene, *ELFN1*. A genetic correlation analysis revealed significant common variant overlaps between RLS and neuroticism (r_g_ = 0.40, se = 0.08, *p*-value = 5.4 × 10^-7^), depression (r_g_ = 0.35, se = 0.06, *p*-value = 2.17 × 10^-8^), and intelligence (r_g_ = -0.20, se = 0.06, *p*-value = 4.0 × 10^-4^). Our study expands the understanding of the genetic architecture of RLS and highlights the contributions of common variants to this prevalent neurological disorder.

## Introduction

Restless legs syndrome (RLS) is a common neurological disease characterized by an irresistible urge to move the legs^1^. Affected individuals exhibit exhaustion and sleepiness, which affect daily activities, work productivity, and personal relationships^2,3^. Studies reported that 5-15% of the European and North American populations suffer from RLS^4,5^, leading to a substantial socio-economic burden. Familial aggregation^6^ and twin studies^7^ estimate its heritability to be approximately 70%, suggesting a major genetic predisposition to RLS. To date, linkage studies in multiplex families have implicated eight genomic regions in families with RLS^8^, and genome-wide association studies (GWASs) have identified an additional 22 genetic risk loci (23 independent variants) associated with RLS^9,10^. Despite this, only about 12% of the heritability is explained^9^, meaning that much remains to be uncovered. Thus far, sequencing studies have been done through the targeted gene approach, and no causal variants were identified within the risk loci^11-13^.

To address this gap and to improve our understanding of RLS’s genetic architecture, we performed a large-scale genomic analysis involving 9,851 cases and 38,957 controls. We identified nine risk loci, of which one has not been previously reported, and performed functional annotations of the detected signals. We also performed pathway and genetic correlation analyses to gain insights into the underlying mechanisms and the relationship between RLS and other traits.

## Methods

### Samples

A total of 9,851 RLS cases and 38,957 controls of European ancestry recruited by three population-based biobanks (CARTaGENE, Canadian Longitudinal Study on Aging, and All of Us) across Canada and the United States were included in the discovery stage meta-analysis of GWAS. Survey-based identification of RLS cases in CARTaGENE and the Canadian Longitudinal Study on Aging was made through essential RLS diagnostic questions. The Personal Medical History domain was used to identify All of Us cases. Controls were selected amongst the individuals with no neurological diseases (Supplementary Table 1).

### Whole genome sequencing and quality assessment

Sequencing was performed by the Genome Centers funded by the All of Us Research Program^14,15^. All centers used the same sequencing protocols that consisted of PCR-free 150 bp, paired-end libraries sequenced on the Illumina NovaSeq 6000 platform and processed using DRAGEN v3.4.12 (Illumina) software. The GRCh38 reference genome was used for alignment^16^. Phenotypic data, ancestry features, and principal components were annotated using Hail through the All of Us Researcher Workbench^17^. Low-quality variants with a call rate of less than 0.95, multiallelic variants, and variants significantly departed from Hardy-Weinberg equilibrium in the control cohort (P ≤ 1.0 × 10^-10^) were removed. Common variants in autosomal chromosomes with a minor allele frequency of higher than 0.005 were included in the association test. Sex concordance was part of the All of Us upstream genomic data quality control process, and all samples within the released genomic data have passed the sex concordance check. Ancestry annotation and relatedness were inferred by the PC-relate method in Hail. Duplicate samples and one of the related participant pairs were excluded^17^.

### Genotyping and imputation

Genotyping data were generated using the Illumina Global Diversity Array for All of Us (1,825,277 markers/GDA-8 v1.0/12,114 participants), Affymetrix protocol for Canadian Longitudinal Study on Aging (794,409 markers/ Axiom 2.0 /29,970 participants), and Illumina Infinium Global Screening Array technology for CARTaGENE (700,078 markers/GSAMD-24v1-0_20011747_A1/2,228 participants). Further cohort and quality control details for the Canadian Longitudinal Study on Aging and CARTaGENE data sets are described in Awadella et al.^18^ and Forgetta et al.^19^. The same quality assessment, filtering, and imputation protocols were applied for all three genotyping cohorts and are summarized in Figure 1. Samples were excluded if missingness was higher than 5% or the reported and genotypic sex was discordant. KING^20^ was used to determine pairwise kinship and ancestry estimation^21^. Unrelated participants with European ancestry were kept for subsequent analyses. Quality assessment for missingness, sex concordance, Hardy-Weinberg equilibrium, and minor allele frequency was conducted using PLINK 2.0^22^. Multiallelic and non-autosomal variants were removed. Variants with a genotyping call rate of less than 99%, a minor allele frequency of less than 0.01, showing nonrandom missingness between cases and controls (P ≤ 1.0 × 10^-4^), and variants that significantly departed from Hardy-Weinberg equilibrium in the control cohort (P ≤ 1.0 × 10^-10^) were excluded.

**Figure 1.**
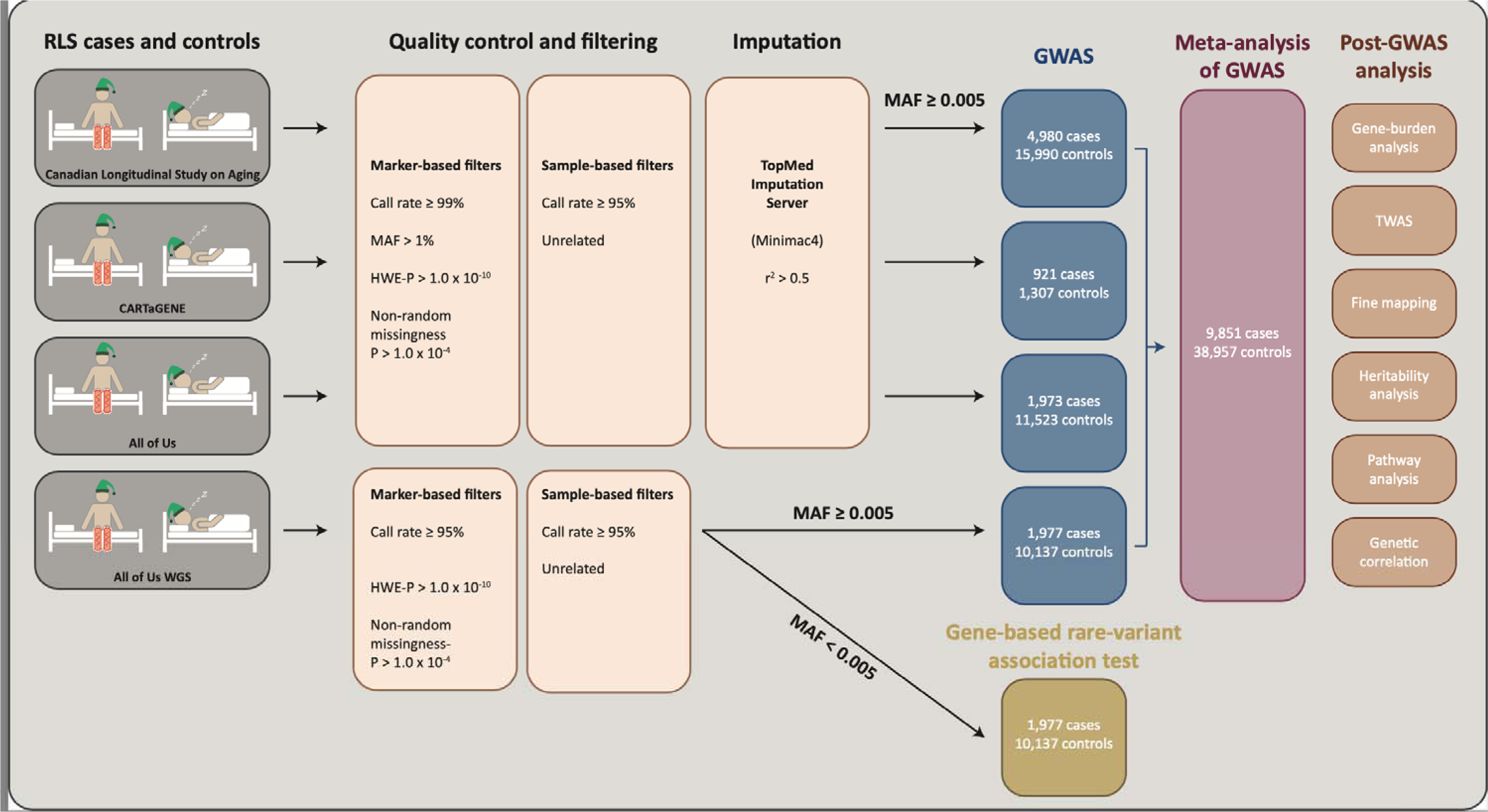
Workflow of the study.

Imputation was carried out using Minimac4 phasing with Eagle v2.4 Trans-Omics for Precision Medicine (TopMed) imputation reference panel (hg38) on TopMed Imputation Server^23-25^. The Canadian Longitudinal Study on Aging cohort was previously imputed via the same pipeline using the TopMed imputation reference panel^19^. After the imputation, variants with a minor allele frequency of less than 0.5% and an imputation quality score of less than 0.5 were excluded prior to the association test.

### Single variant association test and meta-analysis

We generated principal components in PLINK (version 2.0)^22^. We used the step function in the R MASS package to determine the optimum combination of covariates (age, sex, and principal components) to be included in the association tests^26^ (Supplementary Table 3). We performed a logistic regression analysis in PLINK v.2.0^22^ separately in all datasets and meta-analyzed the results using an inverse-variance-weighted meta-analysis in METAL^27^. We included only the variants that were present in all datasets in the final meta-analysis and set the Bonferroni threshold for genome-wide significance as 5.0 × 10^−8^. For follow-up replication of the significant associations in this study, we used the summary statistics of the previous GWAS results by Didriksen et al., which included 10,257 cases and 470,725 controls^10^. The heritability explained by variants tested in our meta-analysis was estimated via LD Score Regression v.1.0.1 using the 1000 Genomes Project cohort for linkage disequilibrium and allele frequencies. We used MungeSumstats to perform standardization of association statistics^28^. We conducted a conditional analysis in PLINK v.2.0 to identify potential secondary signals in the *LMX1B* and *BTBD9* loci that may have been missed in the initial GWAS. The conditional analysis was performed by including the index variant at each locus to the covariates of the logistic regression analyses.

### Genome-wide, gene-based association analyses

Gene-based associations with RLS were estimated with SNP2GENE function in FUMA (v.1.5.1)^29^ using the summary statistics from the RLS meta-analysis (n = 9,851 cases and 38,957 controls). First, we annotated genes that contain RLS-associated variants. For gene annotations, we included all variants that are in linkage disequilibrium (r^2^ > 0.6) with the genome-wide significant signals (P < 5 × 10^-8^). We used UK Biobank European population as a reference panel to define the linkage disequilibrium between variants. Second, we performed a gene-burden analysis using MAGMA (v.1.08)^30^ implemented in FUMA^29^.

For the rare variant burden test, we utilized WGS data of 1,977 cases and 10,137 controls in the All of Us biobank. We annotated potentially disruptive variants using the VEP^31^ and its plugin, Loss-Of-Function Transcript Effect Estimator (LOFTEE)^32^. We performed genome-wide, gene-based SKAT-O analysis using RVTEST (v.2.1.0). We incorporated age, sex, PC2, PC3, PC7, and PC10 as covariates determined by the R MASS package.

### Transcriptome-wide association and fine mapping

For transcriptomic imputation, we applied FUSION^33^ (lasso, susie, top1) and S-PrediXcan^34^ (elastic net, mashr). We tested the differential expression of the available gene models in 13 brain tissue panels imputed in GTEx version 8 of European samples (Table 3). To account for the large number of hypotheses tested, we used a Bonferroni correction p-value of 2.10 × 10^-6^ for FUSION (α = 0.05/23,770 genes tested) and a *p*-value of 3.06 × 10^-6^ for S-PrediXcan (α = 0.05/16,355 genes tested). We conducted a fine mapping approach using FOCUS (v.0.802)^35^. We used our summary statistics, a multiple-eQTL database containing FUSION GTEx version 8 weights, and the 1000 Genomes Project dataset as reference LD. FOCUS assigned a posterior probability of causality for each gene. Finally, the genes in the 90%-credible set with a higher posterior probability were prioritized as putatively causal.

**Table 1.** Genome-wide association study results. *P*-values and the odds ratios were identified through a meta-analysis of the summary statistics (9,851 RLS cases and 38,957 controls). The variant with the lowest p-value is listed for each of the nine loci. The Bonferroni threshold for genome-wide significance was defined as 5.0 × 10^−8^. Genes that are in close proximity to the top variants or identified by post-GWAS analyses are listed. Chr, chromosome; EA, effect allele; OA, other allele; *novel GWAS variant; †prioritized by TWAS or fine mapping.

| Discovery meta-analysis |  |  |  |  | Follow up analysis (Didriksen et al.) |  | Joint stage meta-analysis |  |
| --- | --- | --- | --- | --- | --- | --- | --- | --- |
| Chr:position (hg38) | Alleles (OA, EA) | Gene(s) | OR (95% CI) | *p*-value | OR (95% CI) | *p*-value | OR (95% CI) | *p*-value |
| 1:106,650,227 | G, A | *PRMT6*† | 1.12 (1.08–1.15) | $1.50 \times 10^{-11}$ | 1.15 (1.11–1.18) | $1.90 \times 10^{-17}$ | 1.13 (1.11–1.16) | $3.14 \times 10^{-27}$ |
| 2:66,523,432 | G, T | *MEIS1*† | 1.66 (1.56–1.76) | $2.36 \times 10^{-56}$ | 1.89 (1.89–2.00) | $4.50 \times 10^{-100}$ | 1.77 (1.70–1.85) | $4.50 \times 10^{-152}$ |
| 6:38,476,264 | A, G | *BTBD9, GLO1*† | 0.81 (0.78–0.84) | $2.11 \times 10^{-31}$ | 0.77 (0.74–0.80) | $1.55 \times 10^{-50}$ | 0.77 (0.79–0.81) | $2.02 \times 10^{-79}$ |
| 7:1,340,813 | C, G | *UNCX, ELFN1*† | 0.90 (0.87–0.93) | $2.16 \times 10^{-9}$ | 0.90 (0.87–0.93) | $9.50 \times 10^{-10}$ | 0.90 (0.88–0.92) | $1.17 \times 10^{-17}$ |
| 7:88,682,938 | A, G | *ZNF804B, STEAP2*† | 1.15 (1.10–1.21) | $3.13 \times 10^{-9}$ | 1.13 (1.08–1.18) | $1.69 \times 10^{-7}$ | 1.14 (1.10–1.18) | $3.20 \times 10^{-15}$ |
| 9:8,822,069 | G, A | *PTPRD, PTPRD-AS1*† | 0.91 (0.88–0.94) | $3.36 \times 10^{-8}$ | 0.92 (0.89–0.95) | $9.05 \times 10^{-8}$ | 0.92 (0.90–0.94) | $1.60 \times 10^{-14}$ |
| 9:126,755,162* | C, A | *LMX1B* | 1.14 (1.09–1.19) | $2.20 \times 10^{-9}$ | 1.10 (1.05–1.14) | $2.60 \times 10^{-5}$ | 1.12 (1.08–1.15) | $6.41 \times 10^{-13}$ |
| 15:67,816,234 | C, T | *SKOR1*†, *MAP2K5*† | 0.86 (0.83–0.89) | $1.04 \times 10^{-16}$ | 0.83 (0.80–0.86) | $2.02 \times 10^{-28}$ | 0.84 (0.82–0.87) | $8.73 \times 10^{-43}$ |
| 16:52,610,538 | A, C | *TOX3* | 0.86 (0.83–0.89) | $1.18 \times 10^{-19}$ | 0.82 (0.80–0.85) | $5.88 \times 10^{-34}$ | 0.84 (0.82–0.86) | $4.41 \times 10^{-51}$ |

**Table 2.** Gene-based association test results. Genome-wide gene-based test was done using MAGMA based on the meta-analysis of GWAS discovery cohorts (9,851 RLS cases and 38,957 controls). Bonferroni threshold for genome-wide significance was defined as 2.69 × 10^−6^. The chromosomal position is shown according to hg38.

| Chr | Start | Stop | Gene | *p*-value |
| --- | --- | --- | --- | --- |
| 2 | 66,660,584 | 66,801,001 | *MEIS1* | $1.27 \times 10^{-12}$ |
| 6 | 38,136,227 | 38,607,924 | *BTBD9* | $3.79 \times 10^{-17}$ |
| 6 | 38,643,701 | 38,670,917 | *GLO1* | $8.45 \times 10^{-7}$ |
| 15 | 68,112,042 | 68,126,899 | *SKOR1* | $6.37 \times 10^{-12}$ |
| 15 | 67,835,047 | 68,099,461 | *MAP2K5* | $3.84 \times 10^{-10}$ |
| 16 | 52,471,917 | 52,581,714 | *TOX3* | $5.77 \times 10^{-13}$ |

**Table 3.** Transcriptome-wide association study results. TWAS was performed using FUSION and S-PrediXcan based on the meta-analysis of GWAS discovery cohorts (9,851 RLS cases and 38,957 controls). Bonferroni correction threshold was defined as 2.10 × 10^−6^ for FUSION and 3.06 × 10^−6^ for S-PrediXcan. The chromosomal positions are shown according to hg38.

| Tissue | Gene | Chr | Start | End | Model | TWAS Z-score | TWAS $p$-value | Software |
| --- | --- | --- | --- | --- | --- | --- | --- | --- |
| cortex | *MEIS1* | 2 | 66,433,452 | 66,573,869 | elastic net | 4.67 | $3.05 \times 10^{-6}$ | S-PrediXcan |
| caudate (basal ganglia) | *GLO1* | 6 | 38,703,140 | 38,703,141 | susie | -5.51 | $3.68 \times 10^{-8}$ | FUSION |
| cerebellar hemisphere | *GLO1* | 6 | 38,703,140 | 38,703,141 | lasso | -5.15 | $2.64 \times 10^{-7}$ | FUSION |
| anterior cingulate cortex | *GLO1* | 6 | 38,703,140 | 38,703,141 | mashr | -5.54 | $3.01 \times 10^{-8}$ | S-PrediXcan |
| caudate (basal ganglia) | *GLO1* | 6 | 38,703,140 | 38,703,141 | mashr | -4.84 | $1.28 \times 10^{-6}$ | S-PrediXcan |
| caudate (basal ganglia) | *GLO1* | 6 | 38,703,140 | 38,703,141 | elastic net | -4.81 | $1.49 \times 10^{-6}$ | S-PrediXcan |
| cerebellar hemisphere | *GLO1* | 6 | 38,703,140 | 38,703,141 | elastic net | -5.33 | $9.97 \times 10^{-8}$ | S-PrediXcan |
| hippocampus | *GLO1* | 6 | 38,703,140 | 38,703,141 | mashr | -5.40 | $6.75 \times 10^{-8}$ | S-PrediXcan |
| hypothalamus | *GLO1* | 6 | 38,703,140 | 38,703,141 | mashr | -5.39 | $6.94 \times 10^{-8}$ | S-PrediXcan |
| nucleus accumbens | *GLO1* | 6 | 38,703,140 | 38,703,141 | elastic net | -5.50 | $3.76 \times 10^{-8}$ | S-PrediXcan |
| nucleus accumbens | *GLO1* | 6 | 38,703,140 | 38,703,141 | mashr | -5.40 | $6.59 \times 10^{-8}$ | S-PrediXcan |
| pituitary | *GLO1* | 6 | 38,703,140 | 38,703,141 | elastic net | -4.69 | $2.69 \times 10^{-6}$ | S-PrediXcan |
| putamen (basal ganglia) | *GLO1* | 6 | 38,703,140 | 38,703,141 | elastic net | -4.67 | $2.97 \times 10^{-6}$ | S-PrediXcan |
| substantia nigra | *GLO1* | 6 | 38,703,140 | 38,703,141 | mashr | -5.31 | $1.10 \times 10^{-7}$ | S-PrediXcan |
| hippocampus | *ELFN1* | 7 | 1,688,118 | 1,688,119 | lasso | 5.01 | $5.55 \times 10^{-7}$ | FUSION |
| spinal cord | *UBASH3B* | 11 | 122,655,675 | 122,814,473 | mashr | -4.75 | $2.07 \times 10^{-6}$ | S-PrediXcan |
| anterior cingulate cortex | *MAP2K5* | 15 | 67,542,708 | 67,542,709 | lasso | -5.99 | $2.12 \times 10^{-9}$ | FUSION |
| amygdala | *SKOR1* | 15 | 67,819,703 | 67,819,704 | lasso | 6.03 | $1.62 \times 10^{-9}$ | FUSION |
| frontal cortex | *SKOR1* | 15 | 67,819,703 | 67,819,704 | lasso | 5.68 | $1.37 \times 10^{-8}$ | FUSION |
| hippocampus | *SKOR1* | 15 | 67,819,703 | 67,819,704 | mashr | 6.29 | $3.17 \times 10^{-10}$ | S-PrediXcan |
| putamen (basal ganglia) | *CAPNS1* | 19 | 36,139,574 | 36,139,575 | susie | -5.69 | $1.27 \times 10^{-8}$ | FUSION |

### Regulome-wide association study

RWAS was conducted using the linear model implemented in MAGMA (v.1.10)^36^. Enhancer annotations for eight brain regions (hippocampus, dorsolateral prefrontal cortex, angular gyrus, anterior caudate, cingulate gyrus, inferior temporal lobe, substantial migration, and germinal matrix) were downloaded from the psychENCODE consortium (http://resource.psychencode.org/) and updated in GRCh38 assembly.

### Gene-set enrichment and pathway analysis

A gene-set enrichment analysis approach was conducted using public datasets containing GO (http://geneontology.org) and Reactome (https://reactome.org) pathways in EnrichR^37,38^.

### Genetic correlation

We assessed the genetic correlation between RLS and the following neurological or neuropsychiatric traits of interest with publicly available summary statistics of meta-analysis: Alzheimer’s disease^39^, attention deficit hyperactivity disorder^40^, depression^41^, insomnia^42^, intelligence^43^, neuroticism^44^, and serum iron and ferritin levels^45^. We used LD score regression^46^ with recommended LD scores from the 1000 Genomes Project. A Bonferroni correction p-value of 0.00625 (α = 0.05/8 traits testes) was defined for the significance threshold.

## Results

### Genome-wide inferences

We performed a meta-analysis of GWAS in a total of 9,851 RLS cases and 38,957 controls of European ancestry. These samples included the ‘All of Us’ whole-genome sequencing cohort (1,977 cases and 10,137 controls), the ‘All of Us’ genotyping cohort (1,973 cases and 11,523 controls), the Canadian Longitudinal Study on Aging (4,980 cases and 15,990 controls)^47^, and CARTaGENE (921 cases and 1,307 controls) (Figure 1, Supplementary Table 1). After imputation, quality assessment, and filtering, we performed a GWAS of RLS based on 7,510,495 variants. The estimated sample-size-adjusted genome-wide inflation factor (λ_1,000)_ was 1.0036, indicating minimum residual population structure and confounding.

We identified nine genetic loci that achieved genome-wide significance. Eight of these loci have been previously reported, and one was novel (Table 1). Annotation of the significant variants is provided in Supplementary Table 2. The index variant at this novel RLS-locus was close to the gene *LMX1B* on chromosome 9q33.3 (rs35196838, *p*-value = 2.2 × 10^-9^, OR = 1.14, 95% CI = 1.09–1.19). A conditional analysis did not reveal a secondary signal at this locus (see Supplementary Figure 1 for the regional association and conditional association plots). We replicated the *LMX1B* (rs35196838) variant in an independent cohort of 10,257 cases and 470,725 controls (rs35196838, *p*-value = 2.6 × 10^-5^, OR = 1.10, 95% CI = 1.05–1.14) (Table 1).

Variants in eight known RLS loci exceeded genome-wide significance in our discovery meta-analysis. Nine out of seventeen previously reported risk variants were replicated at Bonferroni significance (*p*-value < 0.05/17 variants = 2.94 × 10^-3^) (Supplementary Table 4). The variant-based heritability was estimated to be 10.17%, whereas the nine significant loci explained 2.16% of the phenotype (21% of the total SNP heritability).

The *TOX3* locus (16q12.1) identified in our RLS meta-analysis is also significantly associated with Parkinson’s disease risk (rs3104783, *p*-value = 1.29 × 10^-12^, OR = 1.07, 95% CI = 1.05–1.09)^48^. However, the direction of the association of this variant in RLS and Parkinson’s disease is different^49^. To further explore the involvement of *TOX3* variants in these two diseases, we created a beta-beta plot and identified a negative correlation coefficient of -0.95 and a Pearson correlation R^2^ of 0.91 (Supplementary Figure 2).

### Gene-burden analysis

To identify genes with variants driving the risk of RLS, we performed a gene-based association analysis using the results of the GWAS meta-analysis using MAGMA (v.1.08)^30^, as implemented in FUMA^29^. We found six significant gene-level associations that achieved genome-wide significance (< 0.05/18,623 protein-coding genes tested = 2.69 × 10^−6^). The strongest of these signals was *BTBD9* (p-value = 3.79 × 10^-17^), followed by *TOX3* (p-value = 5.77 × 10^-13^), both of which are well-established RLS risk loci^9,50,51^. In addition to these GWAS risk genes, we showed that *GLO1*, near the *BTBD9* locus, was significantly associated with RLS (p-value = 8.45 × 10^-7^) (Figure 2b). A conditional analysis showed only one signal near *BTBD9* and *GLO1* (see Supplementary Figure 1c for the regional association plot and Supplementary Figure 1d for the conditional association analysis).

**Figure 2.**
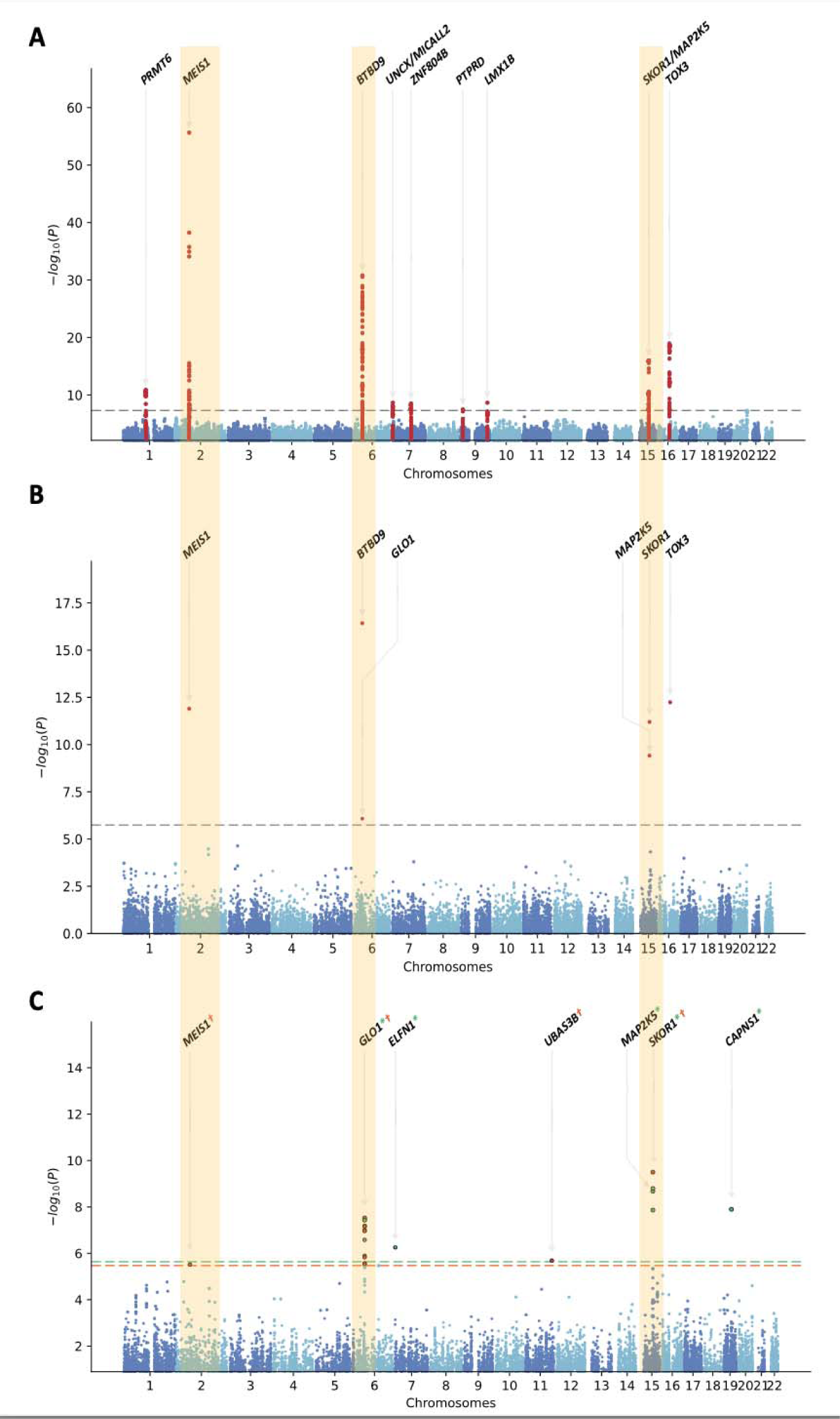
**a.** Manhattan plot for the RLS GWAS discovery cohort (n = 9,851 cases and 38,957 controls; λ_1000_= 1.004), **b.** gene-based common variant analysis using MAGMA (Bonferroni correction threshold = 2.69lJ×lJ10^−6^), and **c.** gene-based transcriptome-wide association study results. Bonferroni threshold was set as 2.10 × 10^-6^ for FUSION and 3.06 × 10^-6^ for S-PrediXcan. Significant genes were labeled green (FUSION) and orange (S-PrediXcan).

We also employed a rare variant burden test to investigate the contribution of rare variants in RLS. Following quality control of sequencing data and annotations, 21,335 genes were tested for rare variant burden in 1,977 cases and 10,137 controls. We subset the rare variants (MAF < 0.005) into high-confidence (such as stop gain or loss, frameshifts, and splice donor or acceptors) and moderate groups (such as missense, inframe insertions, or deletions) based on their calculated variant consequences by VEP^31^. Considering the possible regulatory role of intronic variants, we also conducted the burden test to a group of all rare intragenic variants regardless of their annotation. Our gene-based burden tests did not identify enrichment for rare intragenic single nucleotide variants.

Transcriptome-wide association study identifies new genes associated with RLS.

We sought to integrate expression quantitative trait locus (eQTL) analyses using publicly available transcriptomic data with summary-level GWAS results. To do so, we used two transcriptomic imputation approaches, FUSION^33^ and S-PrediXcan^34^, and tested the differential expression of these gene models in 13 brain tissues (Table 3). We identified 21 associations for seven genes at the transcriptome-wide significant level (Figure 2c), of which four were not identified in the previous transcriptome-wide association study (TWAS) for RLS: *GLO1* (6p21), *ELFN1* (7p22.3), *UBASH3B* (11q24.1), and *CAPNS1* (19q13)^52^. Consistent with previous findings, the expression of three known RLS genes, *MEIS1*, *SKOR1*, and *MAP2K5,* were also associated with RLS^52-54^. A fine-mapping analysis using the ‘Fine mapping Of CaUsal gene Sets’ (FOCUS) software (v.0.802)^35^ prioritized *SKOR1*, *PTPRD-AS1*, *PRMT6*, *STEAP2*, and *GTPBP10* as putative candidate genes for RLS with posterior probabilities of 0.69, 0.13, 0.06, 0.06, and 0.02, respectively (Supplementary Table 6).

Regulatory regions are associated with RLS.

We performed a regulome-wide association study (RWAS) using MAGMA^36^ to identify tissue-specific enhancer and promoter regions associated with RLS. After multiple test corrections for the number of regulatory regions tested in each tissue, we identified significant enrichments of the regulatory regions in *MEIS1, BTBD9, GLO1, PTPRD, MAP2K5, CASC16,* and *TOX3* (See Table 4 for the most significant enrichment for each gene and Supplementary Table 6 for all associations).

**Table 4.** Regulome-wide association study. RWAS was performed using MAGMA based on the meta-analysis of GWAS discovery cohorts (9,851 RLS cases and 38,957 controls). The chromosomal positions are shown according to hg38.

| Tissue | Chr | Start | Stop | Z | p-value | Gene |
| --- | --- | --- | --- | --- | --- | --- |
| Brain dorsotemporal prefrontal cortex | 2 | 66536368 | 66537368 | 7.76 | $4.11 \times 10^{-15}$ | *MEIS1* |
| Brain inferior temporal lobe | 6 | 38374324 | 38375324 | 8.74 | $1.13 \times 10^{-18}$ | *BTBD9* |
| Brain hippocampus middle | 6 | 38701324 | 38702324 | 5.40 | $3.40 \times 10^{-8}$ | *GLO1* |
| Brain angular gyrus | 9 | 8820900 | 8821900 | 5.06 | $2.05 \times 10^{-7}$ | *PTPRD* |
| Brain substantia nigra | 15 | 67802662 | 67803662 | 8.19 | $1.28 \times 10^{-16}$ | *MAP2K5* |
| Brain anterior caudate | 16 | 52592188 | 52593188 | 8.91 | $2.66 \times 10^{-19}$ | *CASC16* |
| Brain germinal matrix | 16 | 52524888 | 52525888 | 5.48 | $2.15 \times 10^{-8}$ | *TOX3* |

**Table 5.** Genetic correlation analysis. The genome-wide genetic correlation was calculated using the LD score regression. rG refers to the genetic correlation between two traits, and SE is the standard error of the genetic correlation. Bonferroni correction threshold was defined as 0.00625 (α = 0.05/8).

| Trait | rG | SE | p-value |
| --- | --- | --- | --- |
| Neuroticism | 0.40 | 0.08 | $5.4 \times 10^{-7}$ |
| Depression | 0.35 | 0.06 | $2.17 \times 10^{-8}$ |
| Intelligence | -0.20 | 0.006 | $4.0 \times 10^{-4}$ |
| Alzheimer's disease | -0.13 | 0.1 | 0.18 |
| Serum iron | -0.09 | 0.07 | 0.20 |
| Serum ferritin | -0.04 | 0.05 | 0.37 |
| Insomnia | 0.03 | 0.04 | 0.55 |
| Attention deficit hyperactivity disorder | 0.03 | 0.05 | 0.56 |

### Gene-set enrichment and pathway analysis

We conducted a gene-set enrichment analysis using Reactome (https://reactome.org) and Gene Ontology (GO; http://geneontology.org) to identify biological pathways associated with the annotated genes. We identified several relevant gene sets including pathways related to neuron differentiation (GO:0030182, *PTPRD*; *SALL1*; *IRX5*; *IRX6*; *PBX3*; and *LMX1B*), generation of neurons (GO:0048699, *PTPRD*; *IRX5*; *IRX6*; *LMX1B*; *MDGA1*) and regulation of myeloid cell differentiation (GO:0045637, *PRMT6*; *MEIS1*) (Supplementary Table 7).

### Genetic correlation

We calculated the genome-wide genetic correlation between pairs of traits using the LD score regression method. After correcting for multiple testing (0.05/7 = 0.007), we identified a significant genetic correlation between RLS and neuroticism^44^ (r_g_ = 0.40, se = 0.08, *p*-value = 5.4 × 10^-7^), depression^41^ (r_g_ = 0.35, se = 0.06, *p*-value = 2.17 × 10^-8^), and intelligence^43^ (r_g_ = -0.20, se = 0.06, *p*-value = 4.0 × 10^-4^) (Table 4).

## Discussion

In the current study, we performed a large GWAS using population-based RLS case-control cohorts that had not been previously studied. We identified and replicated a novel genome-wide significant association in the chromosome 9q33.3 region near *LMX1B.* We also replicated the association of eight known RLS loci at a genome-wide significant level consistent with prior studies^9,10^.

*LMX1B* encodes a homeodomain transcription factor involved in the early specification of midbrain dopaminergic neurons and plays a role in neuronal homeostasis in the adult brain^55,56^. Interestingly, Lmx1b has also been shown to have a regulatory role in the autophagic lysosomal degradation pathway and intracellular transport functions, where its dysfunction is associated with Parkinson’s disease pathogenesis^57^. The altered dopaminergic system is one of the pathological elements in RLS as well, and the implication of *LMX1B* in RLS suggests an overlap between RLS and Parkinson’s disease in terms of their pathogenesis. Moreover, *TOX3* locus (16q12.1), one of the first identified risk factors for RLS, was also shown to be significantly associated with Parkinson’s disease^48^. Together with the previous reports, our findings corroborate the involvement of common molecular mechanisms, and further investigations would help unravel the precise pathways through these common genetic risk factors in RLS and Parkinson’s disease.

In addition to a variant-based analysis, we explored the collective effects of common genetic markers within genes as well as integrated transcriptomic data to identify differentially expressed genes in RLS. Our gene-based approaches identified significant associations for several genes, including well-established RLS risk loci such as *MEIS1*, *SKOR1, MAP2K5,* and *TOX3*. Furthermore, previous identification of MEIS1 protein binding sites within *SKOR1* promoter regions and the regulatory effect of MEIS1 on *SKOR1* expression has prompted our investigation into other regulatory elements that may be associated with the risk of RLS^54^. Interestingly, we identified significant associations of enhancers within the known RLS risk genes, including *MEIS1, BTBD9, GLO1, PTPRD, MAP2K5, CASC16,* and *TOX3*. These results highlighted the potential implication of non-coding regulatory regions in RLS.

Both TWAS and MAGMA analyses identified *GLO1*, located near the *BTBD9* locus, which was previously suggested as an RLS risk gene but had not been replicated yet^11,58,59^. Notably, our TWAS detected a significant association of *GLO1* in 13 gene models in 9 tissues. *GLO1* encodes a major catabolic enzyme, glyoxalase-1, which is involved in the detoxification of methylglyoxal^60^; thus, its inhibition results in the accumulation of reactive carbonyl compounds^61,62^. Considering the implication of methylglyoxal in Parkinson’s disease-like phenotypes^63^ as well as its role in Alzheimer’s disease pathogenesis^64^, our results pinpoint methylglyoxal as a potential therapeutic target for RLS.

Our TWAS showed that increased expression of *ELFN1* (in cis with 7p22.3 locus) is associated with RLS. *ELFN1* has relevant implications in the brain^65,66^ and is associated with several neuropsychiatric and neurodevelopmental disorders, including intellectual disability, attention deficit hyperactivity disorder, and epilepsy^66-68^. Elfn1 protein has been shown to play a role in the modulation of synaptic transmission through its trans interaction with the metabotropic glutamate receptors^69^, which serve as therapeutic targets for several neurological diseases, including Alzheimer’s disease, epilepsy, and Parkinson’s disease^70-72^. Therefore, *ELFN1* may be a promising gene target for functional follow-up in RLS. TWAS also identified two previously unreported genes that are not in cis with any of the GWAS risk loci identified, *UBASH3B* and *CAPSN1.* Although trans-eQTLs could be relevant for many complex diseases, their effect is weaker, and this low statistical power yields low replication rates^73,74^. Therefore, association of these genes should be interpreted with caution. Overall, this integration of transcriptomic information provides additional evidence for the involvement of the associated genes in RLS and expands our understanding of the molecular mechanisms underlying the disorder.

Our study has limitations. The limited availability of non-European samples hindered our ability to carry out comprehensive analyses involving multiple ancestral groups. In addition, cases and controls were selected based on self-report, which may result in diagnostic errors. Nevertheless, the replication of previous findings and overall consistency among the three cohorts used in this study suggested that population-based biobanks are able to capture the pertinent genetic patterns.

## Data availability

The GWAS summary statistics generated in this study have been deposited NHGRI-EBI GWAS Catalog at https://www.ebi.ac.uk/gwas/ (accession IDs GCSTXXXXX). Data are available from the Canadian Longitudinal Study on Aging (www.clsa-elcv.ca) for researchers who meet the criteria for access to de-identified CLSA data.

## Author contributions

FA conceived the study, analyzed the data, and wrote the first draft. PR performed the regulome-wide association study. MR assisted with the annotation for the rare-variant association test. RC, SS, JPR, CL, and AR contributed intellectually to data analysis and manuscript editing. PT, BJT, GAR, and SWS conceptualized and supervised the study. All authors critically reviewed and edited the article.

## Acknowledgments

This research was undertaken in part by the Fonds de Recherche du Québec–Santé. J.P.R. has received a Canadian Institutes of Health Research Frederick Banting & Charles Best Canada Graduate Scholarship (FRN 159279). C. L. has received a CIHR Banting Fellowship. G.A.R. holds a Canada Research Chair in Genetics of the Nervous System and the Wilder Penfield Chair in Neurosciences. This work was supported in part by the NIH Intramural Research Program, the US National Institute on Aging (NIA) grant Z01-AG000949-02 and the US National Institute of Neurological Disorders and Stroke (NINDS) grants NS03130 and ZIANS003154. This study has used the high-performance computational capabilities of the Biowulf Linux cluster at the National Institutes of Health, Bethesda, MD, USA (https://biowulf.nih.gov). The All of Us Research Program is supported by the National Institutes of Health, Office of the Director: Regional Medical Centers: 1 OT2 OD026549; 1 OT2 OD026554; 1 OT2 OD026557; 1 OT2 OD026556; 1 OT2 OD026550; 1 OT2 OD 026552; 1 OT2 OD026553; 1 OT2 OD026548; 1 OT2 OD026551; 1 OT2 OD026555; IAA #: AOD 16037; Federally Qualified Health Centers: HHSN 263201600085U; Data and Research Center: 5 U2C OD023196; Biobank: 1 U24 OD023121; The Participant Center: U24 OD023176; Participant Technology Systems Center: 1 U24 OD023163; Communications and Engagement: 3 OT2 OD023205; 3 OT2 OD023206; and Community Partners: 1 OT2 OD025277; 3 OT2 OD025315; 1 OT2 OD025337; 1 OT2 OD025276. In addition, the All of Us Research Program would not be possible without the partnership of its participants. This research was made possible using the data/biospecimens collected by the Canadian Longitudinal Study on Aging (CLSA). Funding for the Canadian Longitudinal Study on Aging (CLSA) is provided by the Government of Canada through the Canadian Institutes of Health Research (CIHR) under grant reference: LSA 94473 and the Canada Foundation for Innovation, as well as the following provinces, Newfoundland, Nova Scotia, Quebec, Ontario, Manitoba, Alberta, and British Columbia. This research has been conducted using the CLSA dataset [Identification of genetic loci associated with restless legs syndrome in the Canadian population, CLSA Comprehensive Follow-up 1 dataset version 3.1, and Comprehensive Baseline dataset version 6.1 and Genome-wide Genetic Data - Version 3.0, under Application Number 2104033]. The CLSA is led by Drs. Parminder Raina, Christina Wolfson and Susan Kirkland. We thank the CARTaGENE participants and team for data collection and assistance. This research has been conducted using data from CARTaGENE under project number 297405 (https://cartagene.qc.ca/en).

## Supplementary information

**Supplementary Table 1.** Demographic characteristics of study samples in GWAS discovery cohort. Diagnostic questions^†^ in surveys and the Personal Medical History* domain were used to identify cases.

| Dataset | Cases<br>(%<br>females) | Controls<br>(% females) | Mean age,<br>cases/controls | Population | RLS identifier questions |
| --- | --- | --- | --- | --- | --- |
| All of Us<br>WGS* | 1,977<br>(74%) | 10,137<br>(75%) | 62 (± 14), 62 (± 15) | United States | Has a doctor or health care provider ever told you that you have or had any of the following brain and nervous system conditions? (Restless legs syndrome) |
| All of Us<br>GDA* | 1,973<br>(73%) | 11,523<br>(70%) | 62 (± 14), 62 (± 15) |  |  |
| CARTaGENE<sup>†</sup> | 921<br>(67%) | 13,07 (60%) | 56 (± 8), 56 (± 8) | Quebec<br>(Canada) | Do you have restless legs syndrome? |
|  |  |  |  |  | Generally, your discomforts are worse...at rest/during activity/no difference/prefer not to answer/do not know |
|  |  |  |  |  | Generally, your discomforts are relieved by... walking or movement/immobility or relaxation/prefer not to answer/do not know |
|  |  |  |  |  | Generally, your discomforts are worse... in the morning/in the afternoon/evening, bedtime, night/no difference/prefer not to answer/do not know |
| Canadian<br>Longitudinal<br>Study on<br>Aging<sup>†</sup> | 4,980<br>(60%) | 15,990<br>(47%) | 63 (± 10), 63 (± 10) | Canada | Do you have, or have you sometimes experienced, a recurrent need or urge to move your legs while sitting or lying down? |
| Total number | 9,851 | 38,957 |  |  |  |

**Supplementary Table 2.** Gene mapping in the RLS risk regions. Annotation was performed in FUMA (v.1.5.1). It is provided as an Excel file.

**Supplementary Table 3.** Adjusted covariates in the logistic regression test. Covariates were selected using the MASS stepwise function. PC, principal component.

| Dataset | List of the covariates |
| --- | --- |
| CARTaGENE | sex, age, PC1, PC2, PC4, PC7, PC10 |
| Canadian Longitudinal Study on Aging | sex, age, PC2, PC6 |
| All of Us | sex, age, PC3, PC8, PC9, PC10 |
| All of Us WGS | sex, age, PC2, PC3, PC7, PC10 |

**Supplementary Table 4.** Replication of other known RLS risk variants in our meta-analysis.

| Chr | Position (hg38) | EA/OA | Closest gene(s) | Direction | OR | P |
| --- | --- | --- | --- | --- | --- | --- |
| 2 | 3,986,856 | G/A | *DCDC2C* | ++++ | 1.08 (1.05–1.12) | $3.03 \times 10^{-6}$ |
| 2 | 158,343,323 | T/C | *CCDC148* | ---- | 0.93 (0.90–0.97) | $3.14 \times 10^{-4}$ |
| 2 | 189,584,800 | T/A | *SLC40A1* | ---- | 0.96 (0.93–0.99) | 0.016 |
| 2 | 67,842,758 | A/C | *C1D* | NA | NA | NA |
| 3 | 3,406,460 | T/A | *CRBN* | ---- | 0.94 (0.90–0.97) | $3.69 \times 10^{-4}$ |
| 3 | 130,816,723 | G/A | *ATP2C1* | ++++ | 1.07 (1.04–1.11) | $6.25 \times 10^{-5}$ |
| 5 | 171,001,975 | T/C | *RANBP17* | NA | NA | NA |
| 6 | 37,522,755 | G/A | *CCDC167* | ++++ | 1.09 (1.05–1.14) | $1.75 \times 10^{-5}$ |
| 9 | 9,290,311 | T/C | *PTPRD, PTPRD-AS1* | ---- | 0.95 (0.91–0.98) | $1.76 \times 10^{-3}$ |
| 11 | 8,313,948 | A/G | *LMO1* | NA | NA | NA |
| 13 | 72,274,018 | T/G | *DACH1* | ++++ | 1.07 (1.03–1.11) | $6.40 \times 10^{-4}$ |
| 15 | 47,068,169 | T/G | *SEMA6D* | ---- | 0.90 (0.85–0.95) | $2.01 \times 10^{-4}$ |
| 15 | 35,916,797 | T/A | *DPH6* | ---- | 0.90 (0.84–0.96) | $3.01 \times 10^{-3}$ |
| 17 | 48,695,414 | A/G | *PRAC1* | ++++ | 1.04 (1.00–1.08) | 0.08 |
| 18 | 44,290,278 | T/C | *SETBP1* | ++++ | 1.05 (1.01–1.09) | 0.01 |
| 18 | 59,943,413 | T/C | *PMAIP1* | NA | NA | NA |
| 20 | 64,164,052 | G/A | *MYT1* | ---- | 0.91 (0.88–0.94) | $1.17 \times 10^{-7}$ |

**Supplementary Table 5.** Causal posterior probabilities for genes in 90%-credible sets for restless legs syndrome transcriptome-wide association study signals. PIP, posterior inclusion probability.

| Gene | Region | Number of genes<br>in the region | Proxy tissue | Method | PIP |
| --- | --- | --- | --- | --- | --- |
| *PRMT6* | 1:105545220-107867043 | 16 | spinal cord | lasso | 0.06 |
| *STEAP2* | 7:88195689-91032469 | 35 | spinal cord | susie | 0.06 |
| *GTPBP10* | 7:88195689-91032469 | 35 | nucleus accumbens | lasso | 0.02 |
| *PTPRD-AS1* | 9:8456299-9166403 | 7 | spinal cord | susie | 0.13 |
| *SKOR1* | 15:66802429-68725660 | 30 | nucleus accumbens | susie | 0.69 |

**Supplementary Table 6.** Regulome-wide association study (RWAS) results. RWAS was done using MAGMA (v.1.06). Bonferroni correction threshold for each tissue was defined based on the number of enhancers tested. It is provided as an Excel file.

**Supplementary Table 7.** Gene sets and pathways identified by EnrichR using GO biological process and Reactome pathway datasets.

**Supplementary Figure 1.**
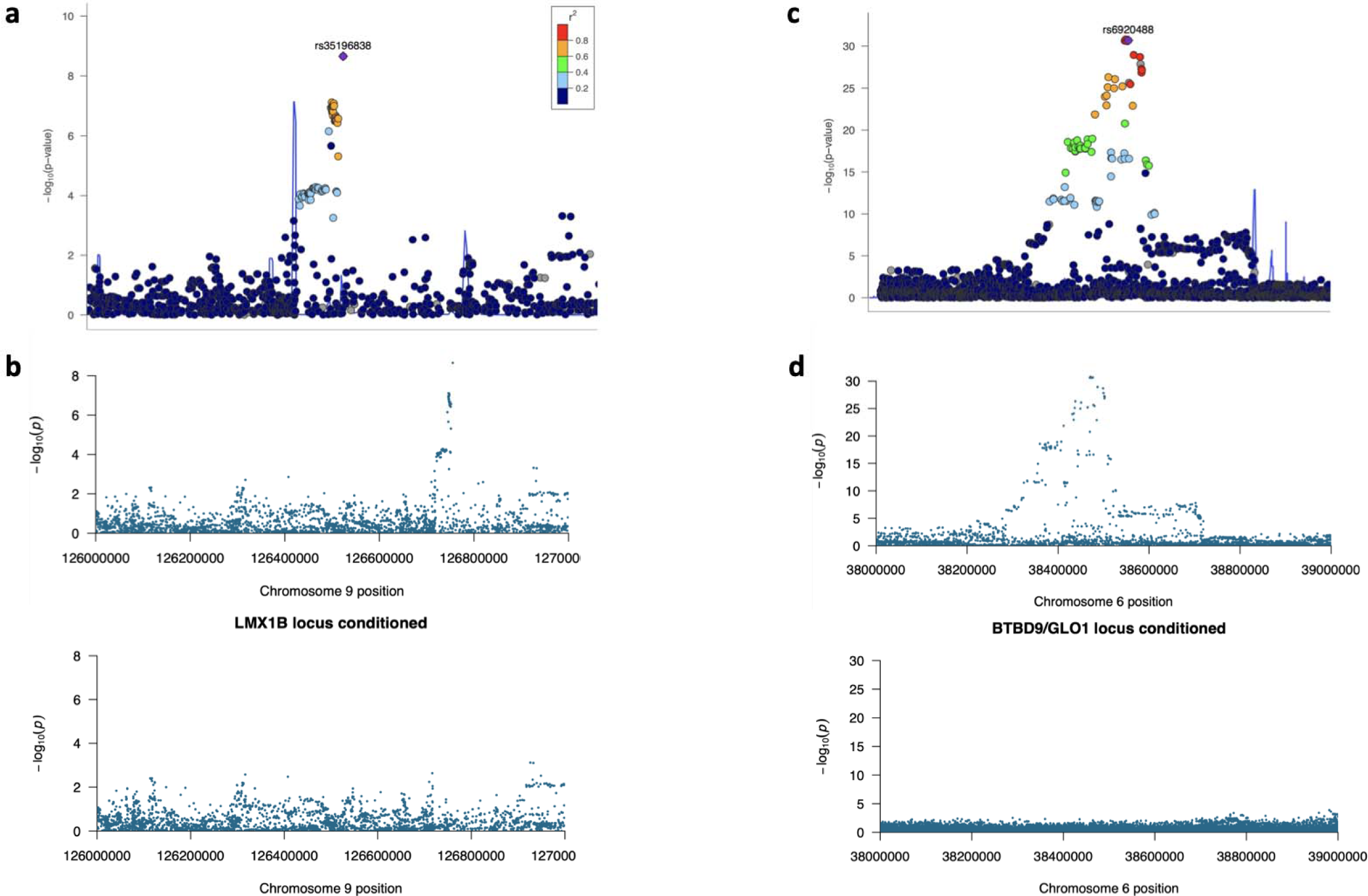
**a.** LocusZoom plot for chr9:126755162:C:A, **b.** Conditional analysis at the *LMX1B* locus (before and after conditioning), **c**. LocusZoom plot for chr6:38476264:A:G, **d.** Conditional analysis at the *BTBD9/GLO1* locus (before and after conditioning).

**Supplementary Figure 2.**
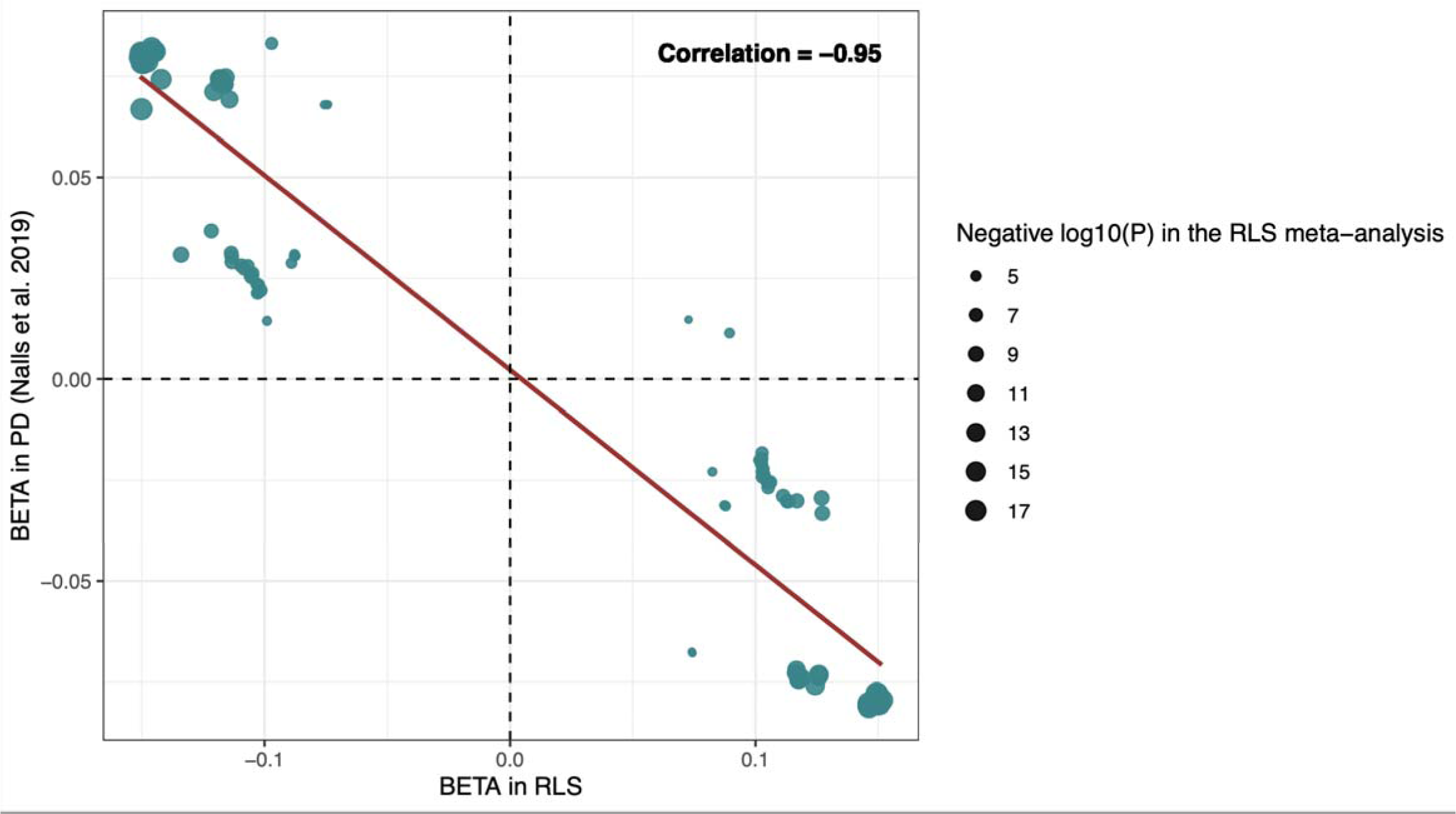
Beta-beta plot for *TOX3* variants in the RLS meta-analysis and Parkinson’s disease meta-analysis. We compared the effect of the *TOX3* variants (P < 1 × 10^-4^) identified in our discovery meta-analysis with the recent Parkinson’s disease GWAS by Nalls et al.. For the top significant variant in Parkinson’s disease (16:52602330:C:A, hg38), we included the beta value from the meta-analysis, whereas, for the rest of the variants, we included summary statistics without 23andMe datasets. A negative correlation was found between the effects of *TOX3* variants in RLS and PD (The correlation coefficient is -0.95, and the Pearson correlation R^2^ is 91).

